# Circulating biomarkers of kidney tumors in TSC (tuberous sclerosis complex) patients

**DOI:** 10.1101/2024.04.01.24305140

**Authors:** Varvara I. Rubtsova, Yujin Chun, Joohwan Kim, Cuauhtemoc B. Ramirez, Sunhee Jung, Elizabeth Cassidy, Gabrielle Rushing, Dean J. Aguiar, Wei Ling Lau, Rebecca S. Ahdoot, Moyra Smith, Sang-Guk Lee, Cholsoon Jang, Gina Lee

## Abstract

Patients with tuberous sclerosis complex (TSC) develop an array of multi-organ disease manifestations, with angiomyolipomas (AML) and cysts in the kidneys being one of the most common and deadly. Early and regular AML/cyst detection and monitoring are vital in lowering TSC patient morbidity and mortality. However, current standard of care for imaging-based methods are not designed for rapid screening, posing challenges for early detection. To identify potential diagnostic screening biomarkers of AML/cysts, we performed global untargeted metabolomics in blood samples from 283 kidney AML/cyst-positive or -negative TSC patients using mass spectrometry. We identified seven highly sensitive chemical features, including octanoic acid, that predict kidney AML/cysts in TSC patients. Patients with elevated octanoic acid have lower levels of very long-chain fatty acids (VLCFAs), suggesting that dysregulated peroxisome activity leads to overproduction of octanoic acid via VLCFA oxidation. This study highlights serum metabolites as novel biomarkers for TSC kidney tumor diagnosis and offers valuable metabolic insights into the disease.

## INTRODUCTION

Tuberous sclerosis complex (TSC) is an autosomal dominant disease that involves neurological manifestations and benign tumors in multiple organs, including the brain, heart, lung, skin, and kidneys (Cook et al., 1996; Lam et al., 2018; Trnka & Kennedy, 2021). TSC occurs in about every 1 in 6000 births, and afflicts about 2 million people worldwide (Henske et al., 2016; Nair et al., 2020; Roach & Sparagana, 2004). TSC most commonly results from de novo spontaneous mutations in *TSC1* or *TSC2* tumor suppressor genes but can also be inherited from parents (Carbonara et al., 1994; Green et al., 1994).

Among TSC-associated manifestations, kidney angiomyolipomas (AML), which are benign tumors composed of blood vessels, muscle, and fat, are the leading cause of death in TSC patients (Eijkemans et al., 2015; Shepherd et al., 1991). AML/cysts develop in 80% of TSC patients during childhood and persistently progress throughout their lives (Ewalt et al., 1998; Kingswood et al., 2019; Rakowski et al., 2006). Treatment of AML/cysts becomes necessary when they become large (>3 cm diameter) and pose a risk for life-threatening rupture and hemorrhage (Bissler & Kingswood, 2016; Cook et al., 1996; O’Callaghan et al., 2004; Rakowski et al., 2006). Therefore, appropriate lifelong surveillance and management of AML/cysts are crucial. Abdominal imaging is recommended at the time of TSC diagnosis since kidney AML can be asymptomatic prior to substantial growth (Hatano & Egawa, 2020; Northrup et al., 2021). Magnetic resonance imaging (MRI) is the preferred modality because it can detect even fat-poor AML with higher accuracy than abdominal ultrasonography or computed tomography (CT). However, definitive diagnosis of AML requires biopsy, which is highly invasive (Buj Pradilla et al., 2017). Further, annual surveillance imaging (Northrup et al., 2021) can pose a financial burden for many patients.

To address these issues and improve the care of AML/cysts in TSC patients, developing an efficient and accessible method of diagnosis is necessary. Such novel biomarkers obtained through simple blood testing would allow physicians to screen for the presence of AML/cysts before performing confirmatory imaging and biopsy-based diagnostics. Modern mass spectrometry-based metabolomics has been used as one of the approaches to discover circulating disease biomarkers (Qiu et al., 2023). To date, there have been minimal metabolomics studies on TSC renal manifestations (Wang et al., 2023). Using global untargeted metabolomics in 283 TSC patients with or without AML/cysts, here we report seven potential blood biomarkers. In particular, the accumulation of octanoic acid in AML/cyst-positive TSC patients provides novel insight into TSC-specific metabolic alterations such as overactive peroxisome-mediated fatty acid oxidation.

## RESULTS

We obtained blood samples from 283 patients with TSC confirmed by clinical diagnostic criteria and genetic testing through the TSC Alliance Biosample Repository and Natural History Database. Patients were divided into two groups, kidney AML/cyst-positive (n=232) or -negative (n=51), based on imaging results from CT, MRI, and ultrasonography. Population characteristics including age, sex, and *TSC* mutation, are shown in Table 1. The majority of AML/cyst-positive patients had a *TSC2* mutation (51.7%) and 10.8% had a *TSC1* mutation, which is consistent with other patient cohorts (Dabora et al., 2001; Rakowski et al., 2006; Sancak et al., 2005). Among the 120 patients with multiple AML lesions, 111 patients have bilateral AML. Other common TSC-related manifestations, including brain subependymal giant cell astrocytoma (SEGA) and cardiac rhabdomyoma, exhibited similar prevalence in the two groups. The use of mTOR inhibitors was also more prevalent in AML/cyst-positive (45.3%) than -negative (23.5%) patients (Table 1).

**Table 1.** Demographic and clinical characteristics of the study population.

|  | TSC with<br>kidney AML or<br>Cysts<br>(N = 232) | TSC with Normal<br>Kidney<br>(N = 51) | P value |
| --- | --- | --- | --- |
| Age in years, median<br>(range) | 16.1 (0.9-58.0) | 13.3 (0.8-52.9) | 0.1015 |
| Gender, n (%) |  |  |  |
| Women | 123 (53.0) | 23 (45.1) | 0.3055 |
| Men | 109 (47.0) | 28 (54.9) |  |
| TSC mutation, n (%) |  |  |  |
| TSC1 mutation | 25 (10.8) | 13 (25.5) | 0.0059 |
| TSC2 mutation | 120 (51.7) | 15 (39.4) |  |
| TSC1&2 mutations | 8 (3.4) | 4 (7.9) |  |
| No | 19 (8.2) | 7 (13.7) |  |
| Unknown | 60 (25.9) | 12 (23.5) |  |
| SEGA, n (%) |  |  |  |
| Yes | 66 (28.4) | 9 (17.7) | 0.1395 |
| No | 155 (66.8) | 37 (72.5) |  |
| Unknown | 11 (4.8) | 5 (9.8) |  |
| Cardiac Rhabdomyoma, n<br>(%) |  |  |  |
| Yes | 120 (51.7) | 25 (49.0) | 0.9224 |
| No | 93 (40.1) | 22 (43.1) |  |
| Unknown | 19 (8.2) | 4 (7.9) |  |
| mTOR inhibitor use, n (%) |  |  |  |
| Yes | 105 (45.3) | 12 (23.5) | 0.0043 |
| No | 127 (54.7) | 39 (76.5) |  |
| Renal manifestation, n (%) |  |  |  |
| Cysts | 90 (38.8) |  |  |
| AML | 32 (13.8) |  |  |
| AML and Cysts | 110 (47.4) |  |  |
| Bilateral AML, n (%) | 112/142 (78.9) |  |  |
| Multiple AML lesions, n<br>(%) | 120/142 (84.5) |  |  |
| AML > 3 cm, n (%) | 44/142 (31.0) |  |  |

To identify potential blood biomarkers of AML/cyst-positive patients, we performed untargeted metabolomics using high-resolution, high-sensitivity liquid chromatography-mass spectrometry (LC-MS). This comprehensive screen yielded 24,070 distinct chemical features, including many unknowns (Figure 1A, B). We selected potential biomarkers by applying selection criteria of |log_2_(fold change)| > 1 and adjusted p<0.05 after false-discovery rate (FDR) correction. A total of 40 chemical features passed these criteria (Table 2). For some of these features, we were able to assign chemical formulas and putative metabolites using accurate mass-to-charge (m/z) values matching publicly available chemical libraries (see Methods). We validated one of the chemical features, m/z =144.11485, as octanoic acid based on m/z and retention time matching with the authenticated chemical standard (Supplementary Fig. 1).

**Figure 1.**
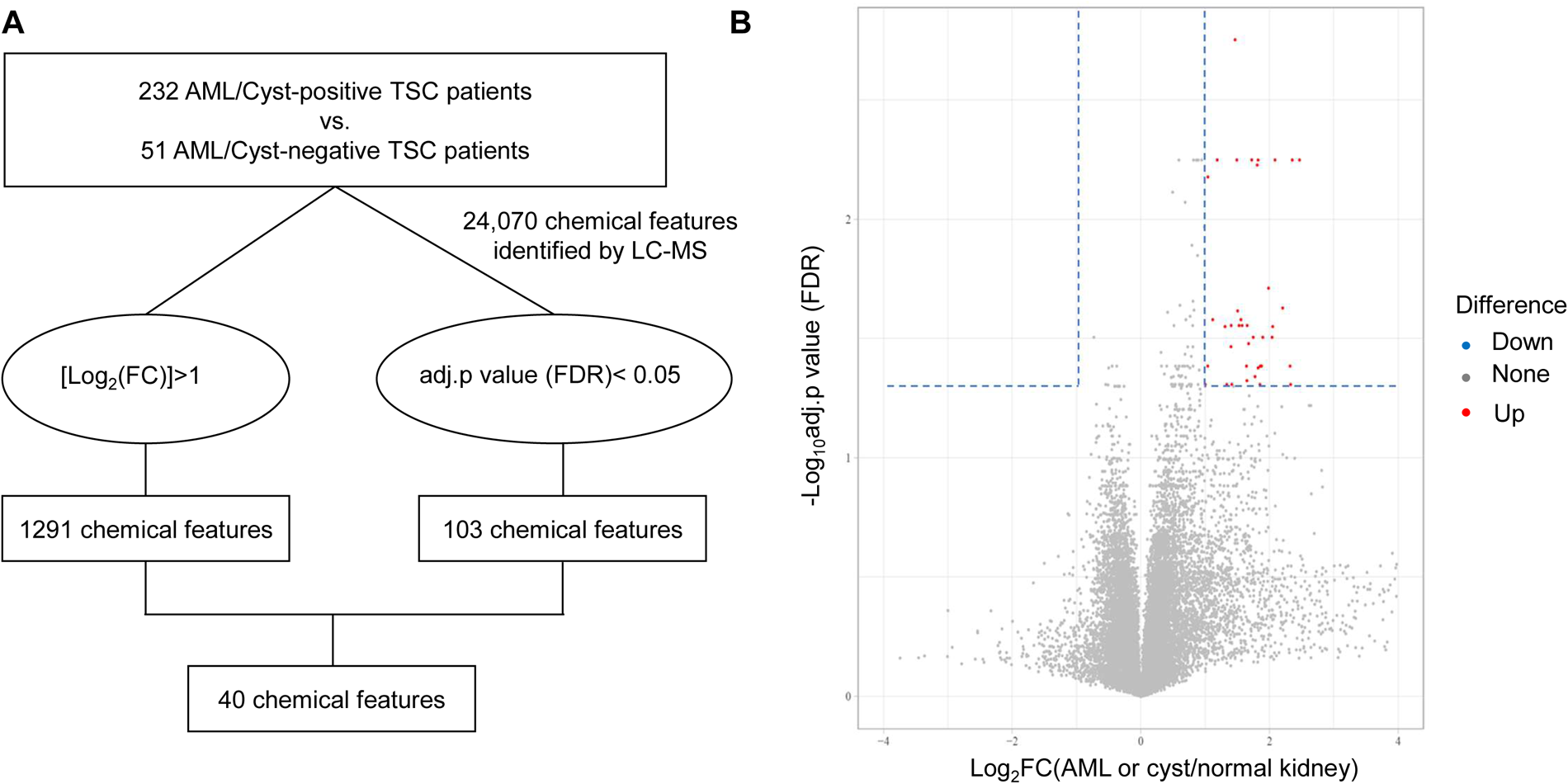
Strategy for metabolomics studies to find plasma chemical features associated with kidney AML/cyst in TSC patients. **A**, Overall schematic to find plasma chemical features associated with kidney AML/cyst. **B**, Volcano plot showing 40 chemical features (red dots) applying selection criteria, |log2(fold change)|>1 and adjusted p-value<0.05.

**Table 2.** Characteristics and subgroups of 40 chemical features monitored by LC-MS.

| Group | MW | RT [min] | Presumed Formula | Presumed metabolite | p value | Adjusted p value | log<sub>2</sub>FC | Max peak area |
| --- | --- | --- | --- | --- | --- | --- | --- | --- |
| 1 | 144.1149 | 2.568 | C8 H16 O2 | Octanoic acid | 0.0001 | 0.042 | 1.823 | 3,629,164,488 |
| 1 | 145.1183 | 2.637 |  |  | <0.0001 | 0.024 | 2.203 | 227,283,883 |
| 1 | 142.0994 | 2.494 | C8 H14 O2 | 2-Octenoic acid | <0.0001 | 0.006 | 2.085 | 135,327,156 |
| 1 | 140.0839 | 2.56 | C8 H12 O2 | (4Z,6Z)-octa-4,6-dienoic acid | <0.0001 | 0.028 | 2.049 | 70,468,118 |
| 1 | 160.1099 | 3.696 | C8 H16 O3 | Hydroxyoctanoic acid | <0.0001 | 0.019 | 1.984 | 59,543,089 |
| 1 | 316.116 | 3.706 | C13 H21 N2 O5 P |  | 0.0001 | 0.041 | 2.319 | 20,759,655 |
| 1 | 144.0599 | 2.609 |  |  | <0.0001 | 0.006 | 2.466 | 15,804,941 |
| 1 | 226.1181 | 2.566 | C8 H14 N6 O2 |  | 0.0001 | 0.046 | 1.775 | 13,716,502 |
| 1 | 146.1214 | 2.568 | C8 H19 P |  | 0.0001 | 0.041 | 1.874 | 11,591,925 |
| 1 | 318.1314 | 3.703 | C13 H23 N2 O5 P |  | 0.0001 | 0.031 | 2.040 | 11,469,040 |
| 1 | 144.1698 | 2.708 |  |  | 0.0002 | 0.049 | 2.328 | 10,854,572 |
| 1 | 308.1986 | 2.602 | C19 H24 N4 |  | <0.0001 | 0.028 | 1.527 | 5,164,287 |
| 1 | 310.2119 | 2.574 | C18 H30 O4 | 13(S)-HpOTrE isomer | 0.0001 | 0.031 | 1.746 | 4,615,159 |
| 1 | 318.1313 | 2.823 | C12 H25 N4 P3 |  | <0.0001 | 0.006 | 2.353 | 3,183,375 |
| 1 | 251.1191 | 2.21 | C10 H21 N O4 S |  | 0.0001 | 0.033 | 1.674 | 3,129,135 |
| 1 | 317.1263 | 2.814 | C18 H15 N5 O |  | 0.0002 | 0.049 | 1.850 | 2,583,266 |
| 1 | 519.3677 | 2.563 | C31 H54 N O P S |  | 0.0001 | 0.041 | 1.871 | 2,165,917 |
| 1 | 72.02022 | 2.542 |  |  | <0.0001 | 0.026 | 1.555 | 1,948,348 |
| 1 | 352.0926 | 3.707 | C13 H26 N2 O P2 S2 |  | <0.0001 | 0.028 | 1.653 | 1,862,380 |
| 1 | 319.1351 | 3.691 |  |  | <0.0001 | 0.028 | 1.574 | 1,489,247 |
| 1 | 354.1081 | 3.688 | C14 H27 O4 P S2 |  | <0.0001 | 0.024 | 1.505 | 1,150,452 |
| 1 | 196.0865 | 3.704 | C7 H17 O4 P |  | <0.0001 | 0.006 | 1.807 | 1,134,695 |
| 1 | 144.1339 | 2.568 |  |  | 0.0001 | 0.031 | 1.895 | 1,042,501 |
| 1 | 224.1023 | 2.5 | C8 H12 N6 O2 | ino-9-(2-hydroxyethoxymethyl)- | 0.0001 | 0.034 | 1.400 | 950,238 |
| 1 | 272.1736 | 3.676 | C13 H24 N2 O4 | N-octanoylglutamine | <0.0001 | 0.028 | 1.405 | 933,994 |
| 1 | 188.0781 | 2.613 | C7 H12 N2 O4 | N-acetyl-glutamine | <0.0001 | 0.006 | 1.723 | 749,064 |
| 1 | 493.0979 | 2.611 | C19 H28 N O8 P S2 |  | 0.0001 | 0.041 | 1.040 | 654,803 |
| 1 | 142.153 | 2.496 |  |  | <0.0001 | 0.006 | 1.822 | 432,102 |
| 1 | 344.7282 | 2.545 |  |  | <0.0001 | 0.026 | 1.116 | 368,752 |
| 1 | 517.3518 | 2.496 | C25 H51 N5 O2 S2 |  | <0.0001 | 0.006 | 1.185 | 217,261 |
| 1 | 144.1058 | 2.494 | C8 H17 P |  | <0.0001 | 0.006 | 1.491 | 195,330 |
| 2 | 211.0819 | 13.339 | C5 H13 N3 O6 |  | 0.0001 | 0.041 | 1.856 | 711,451 |
| 2 | 249.0285 | 13.339 | C4 H9 N7 O2 P2 |  | 0.0002 | 0.048 | 1.649 | 476,283 |
| 2 | 268.0729 | 12.913 | C8 H18 N2 O4 P2 |  | 0.0001 | 0.041 | 1.642 | 422,898 |
| 2 | 351.0831 | 12.049 | C15 H18 N3 O3 P S |  | 0.0002 | 0.049 | 1.414 | 255,705 |
| 3 | 315.24 | 2.972 |  |  | 0.0002 | 0.049 | 1.001 | 11,839,214 |
| 4 | 233.0662 | 1.885 | C6 H12 N5 O3 P |  | <0.0001 | 0.007 | 1.041 | 4,754,931 |
| 5 | 386.0572 | 2.6 | C12 H15 N6 O5 P S |  | 0.0002 | 0.049 | 1.335 | 2,705,805 |
| 6 | 194.0424 | 3.707 | C7 H14 O2 S2 |  | <0.0001 | 0.028 | 1.307 | 1,927,228 |
| 7 | 470.2339 | 2.454 | C19 H40 N2 O7 P2 |  | <0.0001 | 0.002 | 1.464 | 1,320,292 |

We next performed a systematic correlation analysis to understand the biochemical relationships between our identified chemical features. Interestingly, we found significant positive correlations between several chemical features (Supplementary Fig. 2 and 3). Except for only a few cases, most correlations are observed between chemical features that have different retention times, suggesting that they are distinct metabolites in related biochemical pathways. For example, octanoic acid exhibited strong positive correlations with putative 2-octenoic acid (Pearson’s r = 0.922) and hydroxy octanoic acid (Pearson’s r = 0.881) (Figure 2A, B). Based on our systematic correlation analysis, we separated the chemical features into 7 groups (Table 2). The chemical features within each group highly correlate with each other, with a correlation coefficient (r) greater than 0.5 and a p value of less than 0.001. Using this analysis, we sorted 31 chemical features into group 1 (Supplementary Fig. 2) and 4 features into group 2 (Supplementary Fig. 3).

**Figure 2.**
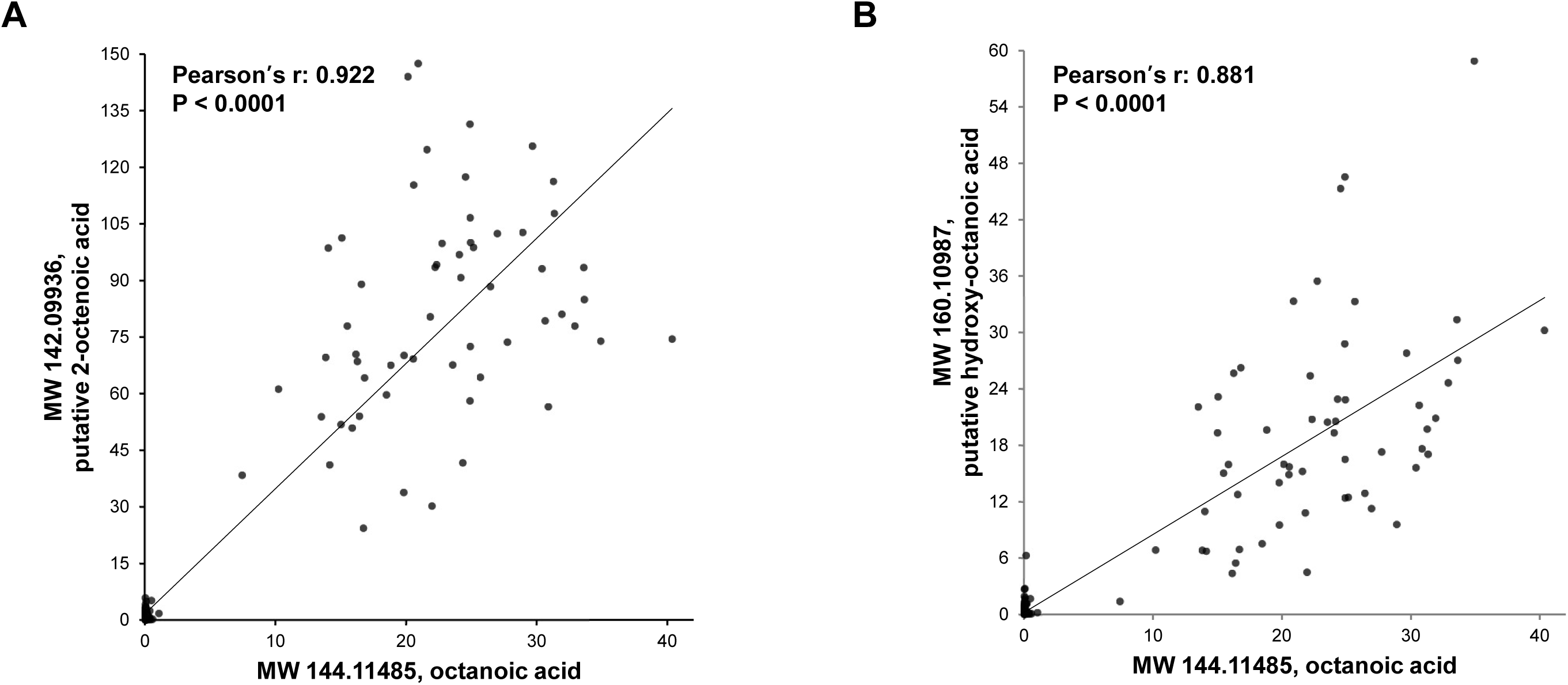
Correlation analysis between the identified chemical features. **A**, Correlation between MW 144.11485 (octanoic acid) and MW 142.09936, (putative 2-octenoic acid). **B**, Correlation between MW 144.11485 (octanoic acid) and MW 160.10987 (putative hydroxy-octanoic acid).

We next obtained 51 non-TSC patients’ blood to examine whether our identified chemical features are specific to AML/cyst-positive TSC patients (Figure 3A-G). For this analysis, we selected the chemical feature with the highest peak area in groups 1 and 2, as well as the individual chemical features that made up groups 3 to 7 (Table 2). Comparison of the abundances of the seven chemical features between the three patient groups (non-TSC vs. AML/cyst-negative TSC vs. -positive TSC) showed significantly higher levels of these chemicals in AML/cyst-positive TSC patients than the other two groups, with octanoic acid level showing the most difference (Figure 3A).

**Figure 3.**
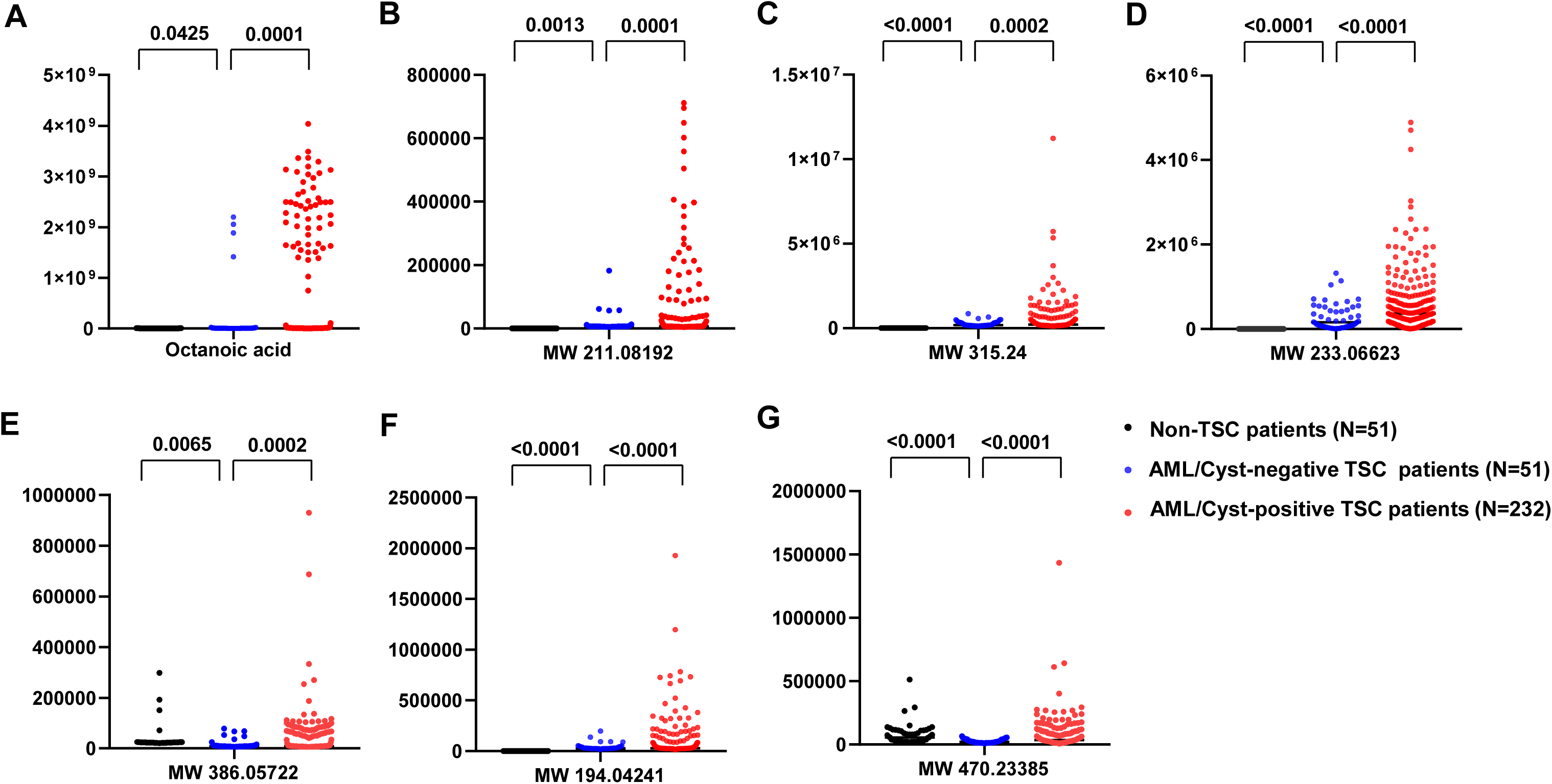
Comparison of abundance of the seven selected chemical features between plasma samples from non-TSC, AML/cyst-negative TSC, and AML/cyst-positive TSC patients. **A-G**, Y-axis indicates ion counts. Note that the abundances of these chemical features are significantly higher in the AML/cyst-positive TSC patients than the other groups, with octanoic acid in **A** showing the greatest differences.

We then evaluated the collective clinical performance of seven chemical features. Following a typical approach to determining biomarker cut-offs (Polley & Dignam, 2021), we considered a blood test positive if any single chemical feature is elevated above the cut-off, which is set at the 90^th^ percentile of AML/cyst-negative TSC patients (Supplementary Table 1). Encouragingly, the frequency of positives (clinical sensitivity) for the diagnosis of AML/cyst according to the increase in one of the seven chemical features was high (80.2%) (Figure 4A). We found AML/cyst-negative patients have a 47.1% frequency of false positives (Figure 4A). These false-positive patients may have small or fat-poor AML/cysts undetected by less sensitive ultrasonography. Also, for some patients, blood was collected several months after the latest imaging-based monitoring, raising the possibility that AML/cysts developed at the time of blood collection. Given that the circulating marker-based method is designed as the initial diagnostic screen prior to confirmative image-based diagnosis (if the blood marker is positive), future studies are warranted to increase the true positive rates to nearly 100%.

**Figure 4.**
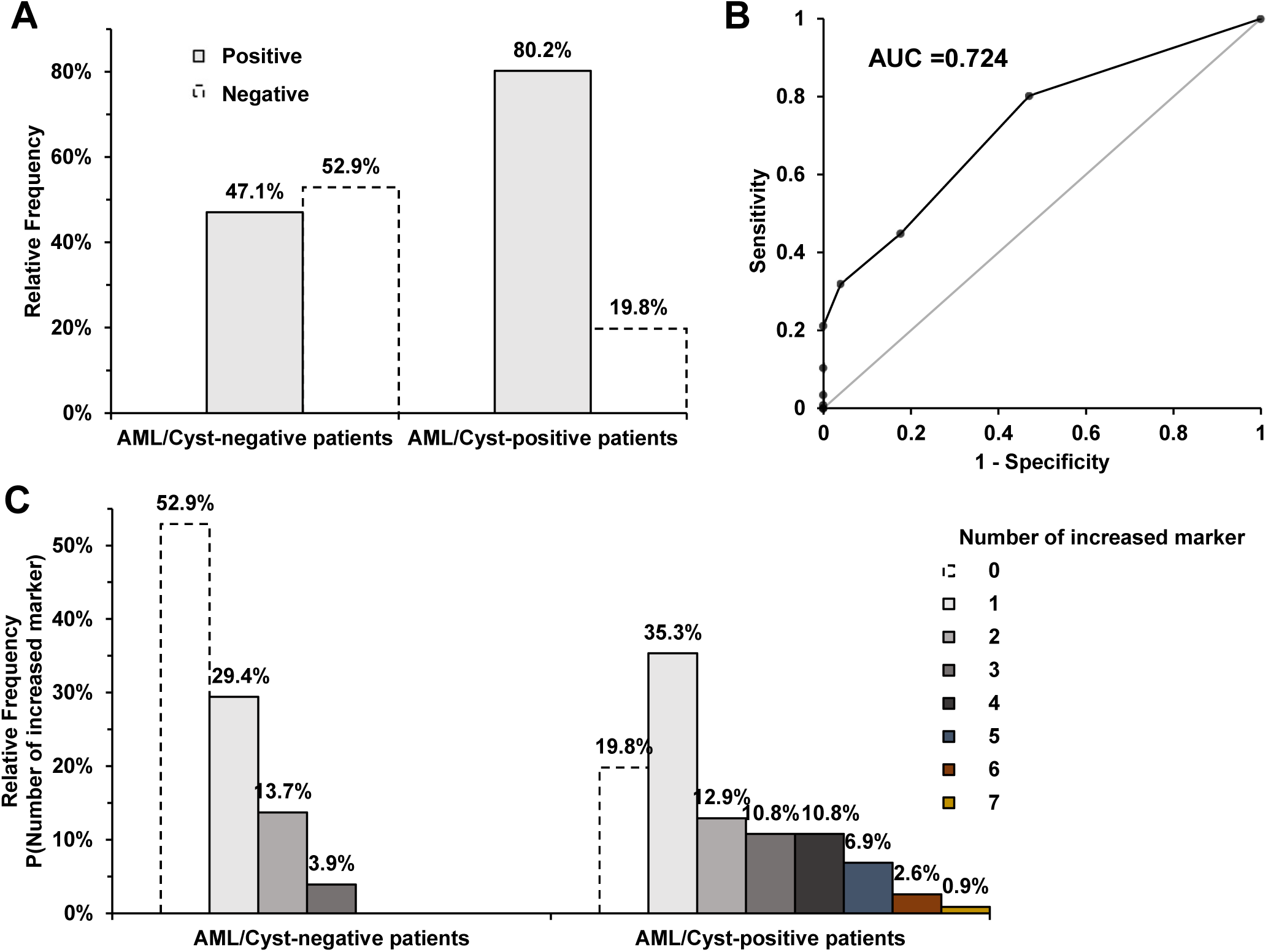
Clinical performance of blood test combining 7 chemical features in detecting AML/cyst. **A**, Relative frequency of AML/cyst-negative vs. -positive patients that are either positive or negative on the test. Positive means one of the 7 chemical features is elevated above the cut-off. **B**, Receiver Operating Characteristic (ROC) curve of the blood test. **C**, Distribution of the number of positive markers in AML/cyst-negative vs -positive patients. Note that no AML/cyst-negative patients showed >3 positive markers. In contrast, the majority of AML/cyst-positive patients showed at least 2 markers positive.

To gauge the sensitivity and specificity of disease biomarker combinations, a Receiver Operating Characteristic (ROC) curve is widely used, with an area under the curve (AUC) above 0.7 being regarded as an acceptable diagnostic marker (Mandrekar, 2010). Importantly, the combination of our 7 chemical features yielded an AUC of 0.724 (Figure 4B), indicating that they can serve as adequate diagnostic markers. Among the combination of seven chemical features, designating positivity as the elevation of any single marker showed the most favorable clinical performance with the highest Youden’s index (Supplementary Table 2). For AML/cyst-negative patients, 29.4% showed one marker positive but the percentile decreased dramatically with more markers, with no patients showing >3 positive markers (Figure 4C). In contrast, the majority of AML/cyst-positive patients showed at least 2 markers positive. This analysis supports the use of combined markers to enhance the sensitivity for AML/cyst detection.

Lastly, our identification of three elevated 8-carbon compounds (octanoic acid, putative 2-octenoic acid, and hydroxy octanoic acid) in a subpopulation of AML/cyst-positive TSC patients (Figure 2) motivated us to scrutinize the underlying biology, which may reflect AML/cyst formation and its influence on kidney metabolism. Firstly, we found that no population characteristics, including age, sex, *TSC* mutation type, mTOR inhibitor treatment, or the presence of other organ disorders, are correlated with the elevation of these 8-carbon compounds (not shown). Secondly, we hypothesized that the rise of 8-carbon compounds is due to a specific diet such as a fat-enriched ketogenic diet frequently used to manage epilepsy in TSC patients (Henske et al., 2016). Indeed, high octanoic acids in blood persisted for a long time in these patients (on average, 843 days apart) (Supplementary Fig. 4 and Supplementary Table 3), supporting the idea that long-term dietary habits may drive their increased levels. However, we found neither medium nor long-chain fatty acids, the main components of ketogenic diet, to be elevated in these patients (Supplementary Fig. 5). Also, 3-hydroxybutyrate, a ketone body, was not elevated (Supplementary Fig. 6). Thus, the select increase of 8-carbon compounds but no other fatty acids or ketones exclude the possibility of simple diet effects.

Aside from diet, octanoic acid can be generated endogenously by peroxisomes, organelles where very long-chain fatty acids (VLCFA) are oxidized into octanoic acid. Octanoic acid is then further oxidized in mitochondria (Lazarow & De Duve, 1976; Reddy & Hashimoto, 2001). In this scenario, elevated octanoic acid may reflect highly active peroxisomal VLCFA oxidation, unmatched by subsequent mitochondrial oxidation. Indeed, patients with high octanoic acid levels had significantly lower VLCFAs in plasma (Figure 5A and Table 3), suggesting their accelerated breakdown. Further, there was a significant negative correlation between octanoic acid and many VLCFAs (Figure 5B and Table 3). Consistently, from RNA-sequencing data of kidney tissues from TSC and normal patients (Martin et al., 2017), we found that the kidneys from TSC patients with AML show highly elevated expression of *peroxisome proliferator-activated receptors gamma* (*PPAR-gamma*), which may mediate VLCFA oxidation (Lu et al., 2022) (Figure 5C). Together, these data suggest that elevated octanoic acid likely reflects hyperactive peroxisomal VLCFA oxidation in the kidneys of TSC patients with AML/cysts (Figure 6).

**Figure 5.**
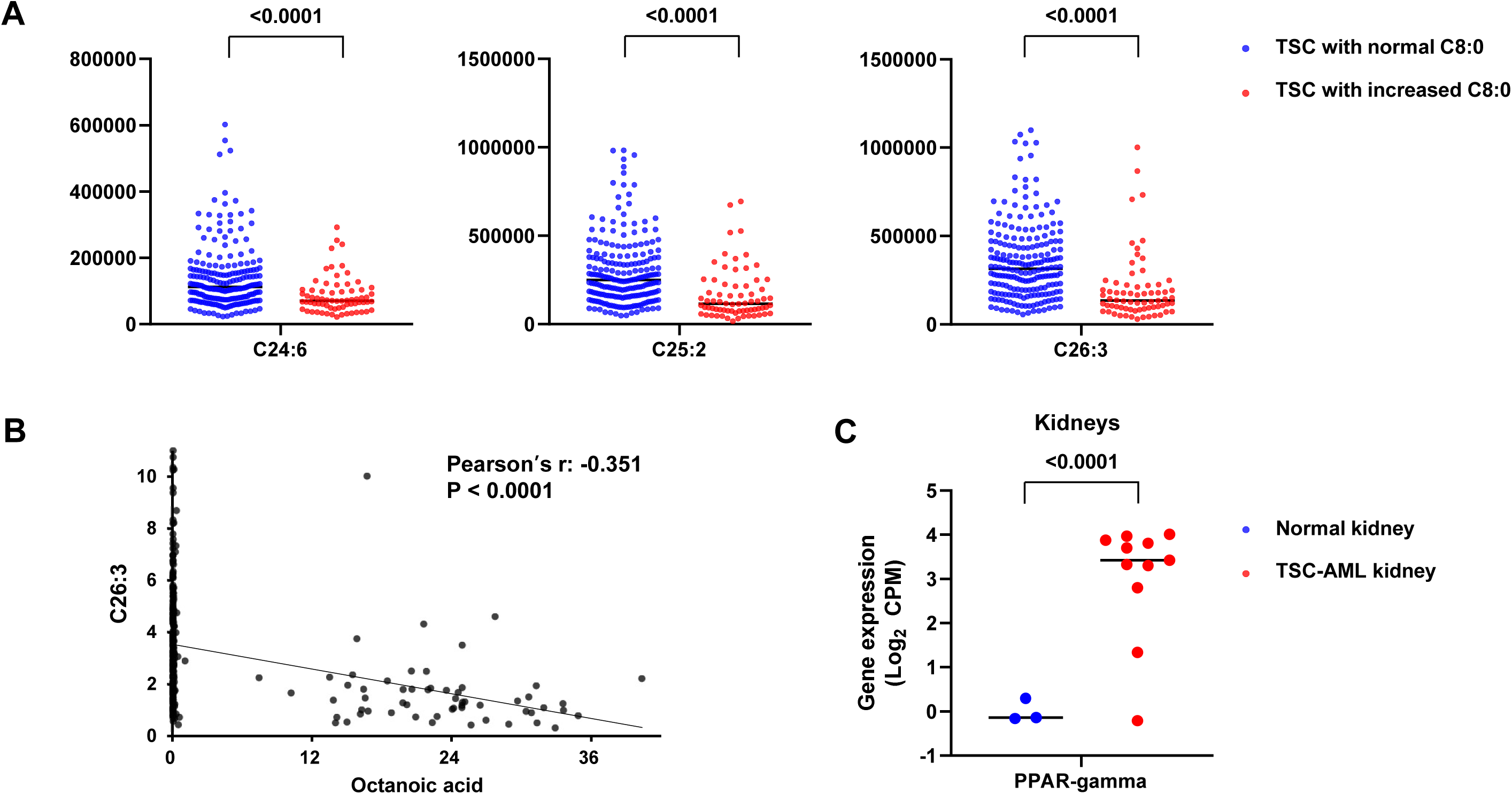
Blood octanoic acid increase is linked to peroxisome activity. **A**, Ion counts of plasma VLCFAs in AML/cyst-positive patients with normal vs. increased octanoic acid. **B**, A significant negative correlation between the abundance of octanoic acid and that of very long-chain fatty acid (VLCFA) C26:3 in AML/cyst-positive patients. **C**, Gene expression of peroxisome proliferator-activated receptors gamma (PPAR-gamma) in normal kidney (n=3) vs TSC-AML kidney (n=11).

**Figure 6.**
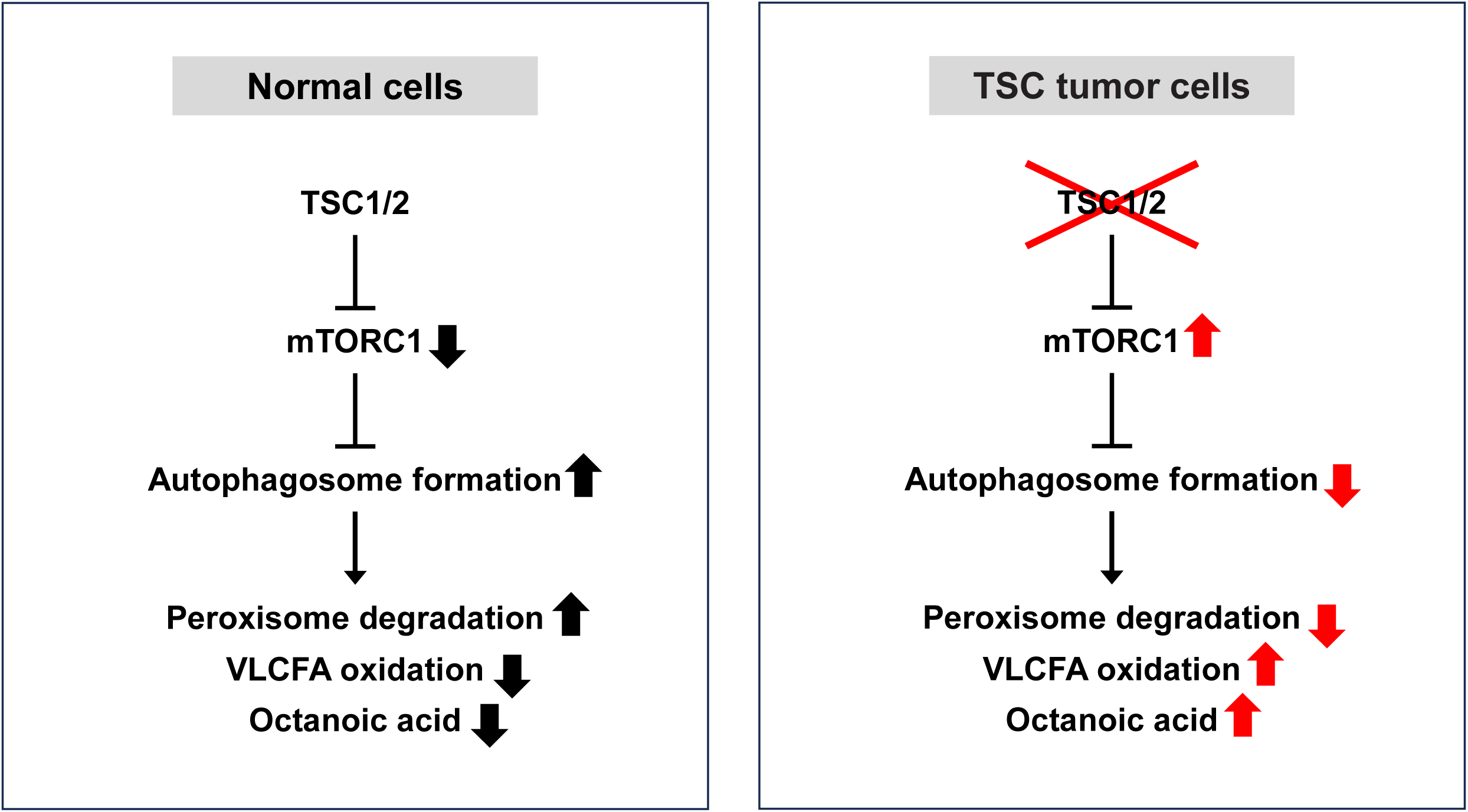
Schematic of increased octanoic acid and decreased very long-chain fatty acids (VLCFA) levels in AML/cyst-positive TSC patients. In the peroxisomes, VLCFAs are oxidized to octanoic acid. In normal cells, the TSC1/2-mTORC1 was shown to degrade peroxisomes by autophagosome formation (Alexander et al., 2010; Zhang et al., 2013). In cells without TSC1/2 activity, mTORC1 is constitutively active and peroxisomes can remain intact and continue to oxidize VLCFAs, which leads to increased production of octanoic acid.

**Table 3.**
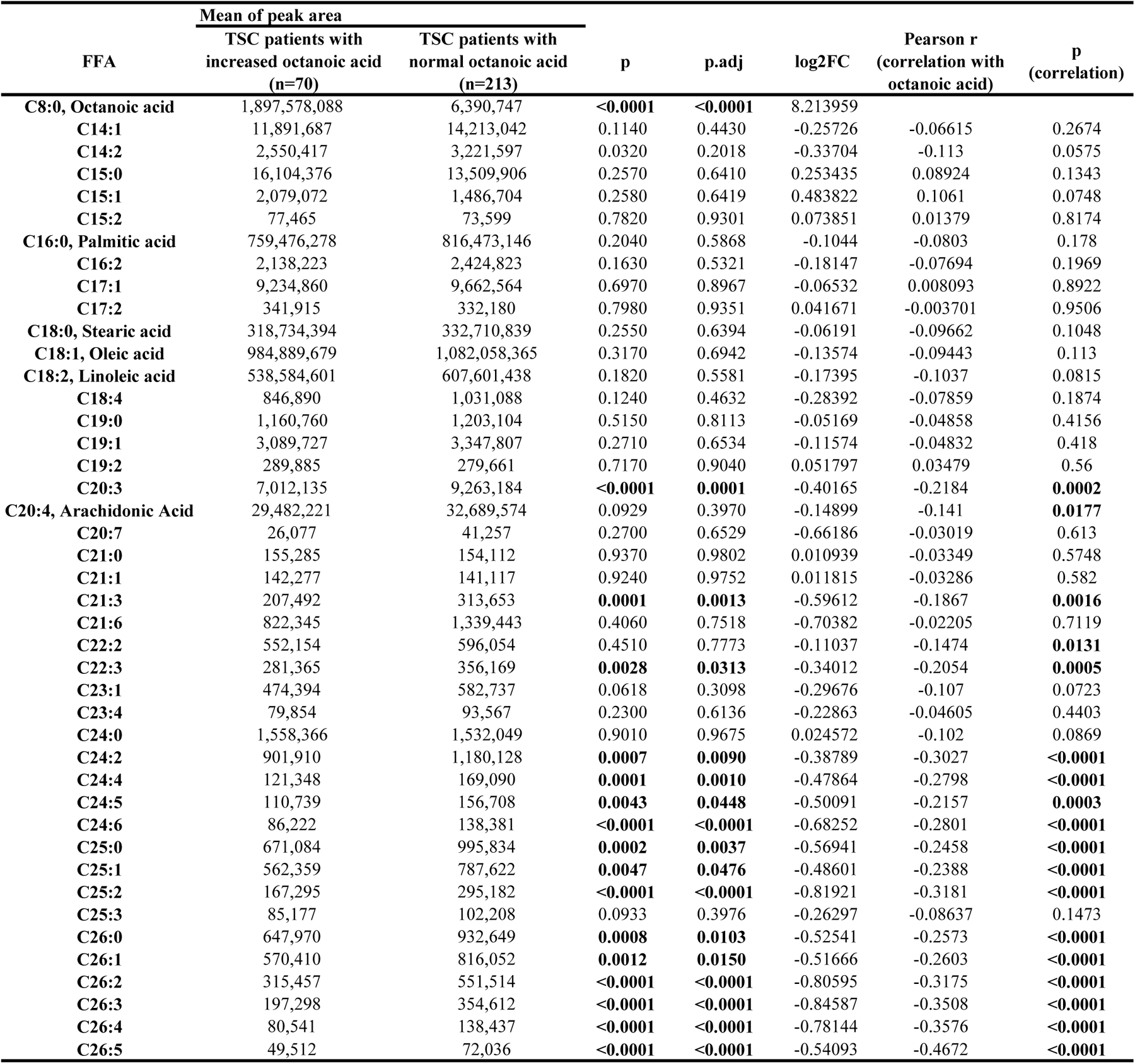
Difference in free fatty acid levels between TSC patients with increased octanoic acid and those with normal octanoic acid.

## DISCUSSION

The challenges associated with diagnosing and monitoring kidney AML or cysts in TSC patients highlight the need to develop simpler, more affordable, and less invasive screening methods. In our analysis of plasma samples from 283 AML/cyst-positive and -negative TSC patients, we identified octanoic acid and six additional chemical features that are significantly increased in AML/cyst-positive TSC patients as compared to AML/cyst-negative TSC patients, regardless of mTOR inhibitor treatment. Using all seven circulating biomarkers combined is a highly promising diagnostic tool for kidney AML or cysts (Figure 4).

With this convenient detection method, physicians can first screen patients with these biomarkers in patients’ blood, and then use standard imaging and biopsy techniques to confirm the presence of kidney AML/cysts. This removes the reliance on imaging and biopsy as the first step in patient evaluation. Of note, a similar approach has been done for the TSC-associated condition lymphangioleiomyomatosis (LAM) (Bottolo et al., 2020; Chang et al., 2012; McCarthy et al., 2021; Nijmeh et al., 2018; Revilla-López et al., 2023; Seyama et al., 2006; Young et al., 2008) where vascular endothelial growth factor-D (VEGF-D) has been identified as a circulating biomarker that can be used toward LAM diagnosis (Seyama et al., 2006; Young et al., 2008). Future studies are required to determine whether our identified markers can determine stage or severity of disease and monitor drug effect (regression of AML/cysts).

The identified diagnostic markers also provide insight into mechanisms by which kidney AML/cysts develop in TSC patients. We noted markedly elevated levels of octanoic acid and closely related metabolites such as putative 2-octenoic acid and hydroxy octanoic acid in a subset of AML/cyst-positive TSC patients (Figure 2). In addition, the strong negative correlation that we observed between these metabolites and several VLCFAs provides potential biological insights that underlie AML/cyst-positive kidneys (Figure 5B and Table 3). VLCFAs are oxidized in the peroxisomes, which are particularly abundant in the liver and kidney (Jo et al., 2020; Litwin et al., 1988; Titorenko & Rachubinski, 2001). During VLCFA oxidation in these organs, the peroxisome produces reactive oxygen species (ROS), which can cause *TSC1* and *TSC2* to associate with peroxin (PEX) proteins on the peroxisomal membrane. This in turn suppresses mTORC1 and triggers peroxisome degradation via autophagy (Alexander et al., 2010; Zhang et al., 2013). Peroxisome abundance is therefore negatively regulated by the TSC1/2-mTORC1 signaling-dependent “pexophagy” (Walker et al., 2018; Zhang et al., 2015). However, when both alleles of *TSC1* or *TSC2* are mutated in AML or cysts, mTORC1 cannot be suppressed and peroxisomes can remain active and continue to oxidize VLCFAs. This can result in an increased production of octanoic acid, which may then be released into the bloodstream (Figure 6). This intriguing connection between *TSC* mutations and increased peroxisome activity may provide a biological explanation for the increased octanoic acid.

## METHODS

### Sample collection and ethics

The Institutional Review Board of Salus IRB Services (formerly known as Ethical & Independent Review Services) and UC Irvine gave ethical approval for this work. All samples and clinical information were collected in compliance with Salus IRB Services Protocol #15039 (TSC Alliance), UC Irvine IRB Protocol #2012-8716, and UC Irvine IRB Protocol #2021-6823 in accordance with the ethical standards of the responsible committee on human experimentation (institutional and national) and with the Helsinki Declaration of 1975, as revised in 2013. Blood samples were collected and provided by the TSC Alliance Biosample Repository (283 TSC patient samples) and UC Irvine Experimental Tissue Resource (51 non-TSC patient samples) with the clinical information provided by the TSC Natural History Database and UC Irvine Health, respectively. For a longitudinal study, blood was collected twice from the same patient (n=51) with a mean interval of 843 days (range of 176-1642 days). Briefly, the blood collected in the ethylenediaminetetraacetic acid (EDTA)-coated vacutainer blood collection tube was centrifugated at 500g at 4°C for 10 min. The clear supernatant (plasma) was centrifugated again at 1,500g at 4°C for 10 min, and the supernatant was stored at -80°C until metabolomics analysis. All TSC subjects had a definite clinical diagnosis of TSC meeting the standard inclusion criteria (Northrup et al., 2021).

### Measurements of metabolites using LC-MS

For aqueous metabolites extraction, serum (5 µL) was mixed with 150µl -20°C 40:40:20 methanol:acetonitrile:water (v:v:v, extraction solvent), vortexed and immediately centrifuged at 16,000g for 10 min at 4°C. The supernatant (70 µL) was loaded into individual LC-MS vials. Metabolites were analyzed by quadrupole-orbitrap mass spectrometer (Q-Exactive Plus Quadrupole-Orbitrap, Thermo Fisher) mass spectrometers coupled to hydrophilic interaction chromatography (HILIC) via electrospray ionization. LC separation was on an Xbridge BEH amide column (2.1 mm x 150 mm, 2.5 µm particle size, 130 Å pore size; Waters) at 25°C using a gradient of solvent A (5% acetonitrile in water with 20 mM ammonium acetate and 20 mM ammonium hydroxide) and solvent B (100% acetonitrile). The flow rate was 350 µL/min. The LC gradient was: 0 min, 75% B; 3 min, 75% B; 4 min, 50% B; 5 min, 10% B; 7 min, 10% B; 7.5 min, 75% B; 11 min, 75% B. Column temperature set to 25°C. Autosampler temperature was set at 4°C and the injection volume of the sample was 5 μL. MS analysis was acquired in negative ion mode with MS Full-scan mode from m/z 70 to 830 and 140,000 resolution. Data analysis was performed with Compound Discoverer and MAVEN software.

### Gene expression analysis

The differentially expressed gene sets of the TSC renal angiomyolipoma (TSC-AML) and normal TSC kidney tissues were adopted from Martin et al., 2017 (Martin et al., 2017). To assess the genes involved in peroxisome biogenesis and fatty acid beta-oxidation, the list of the genes under this functional category was retrieved from the QuickGO database (https://www.ebi.ac.uk/QuickGO/) and used to identify the gene expression changes in between the TSC AML and non-TSC kidneys.

### Statistical analysis

Selection criteria for determining chemical features of interest included: (1) **|**log_2_(fold change)**|** > 1; and (2) demonstration of a statistically significant difference between TSC patient with kidney AML or cysts and those with normal kidney using a t-test with FDR correction. Mann-Whitney test was used to compare median of age between groups. Pearson’s correlation was used to evaluate the correlation between two variables. Comparison of categorical measures between independent groups was performed by Pearson’s chi-squared test. Student’s t-test was used to compare group means of peak area of chemical features and gene expression analysis. Statistical analyses were performed with R, version 4.1.3 (http://www.r-project.org) and Analyse-it (version 6.15, Analyse-it Software, Ltd., Leeds, UK).

## Data Availability

All data produced in the present study are available upon reasonable request to the authors.

## Acknowledgments

We thank all members of the Jang and Lee laboratories for the discussion. We also would like to acknowledge the support of the Chao Family Comprehensive Cancer Center Experimental Tissue Shared Resource Facility. This work was supported by Department of Defense W81XWH2110254 (to G.L.), National Institutes of Health K22CA234399 (to G. L.), TSC Alliance BSR-01-23 (to G.L.), R01AA029124 (to C.J.), and P30CA062203 (University of California Irvine, Chao Family Comprehensive Cancer Center). J.K. was supported by a postdoctoral fellowship from the TSC Alliance (02-23). C.B.R. was supported by predoctoral fellowship from the University of California Irvine, Interdisciplinary Cancer Research program (T32CA009054). S.J. was supported by a postdoctoral fellowship from the National Research Foundation of Korea (2021R1A6A3A14039681). We would also like to thank the TSC Alliance and all contributors to the TSC Natural History Database and Biosample Repository, including the Van Andel Research Institute, which organized, processed, and distributed samples utilized.

## Author contributions

S.L., C.J., and G.L. conceived the project and supervised the study. S.L. performed all the LC-MS running and data analysis. Y.C. performed bioinformatic analysis of RNA-seq results. E.C. and G.R. contributed to sample collection. V.I.R. S.L., C.J., and G.L. wrote the original manuscript. All authors critically reviewed the results and edited the manuscript.

## References

Alexander, A., Cai, S.-L., Kim, J., Nanez, A., Sahin, M., MacLean, K. H., Inoki, K., Guan, K.-L., Shen, J., Person, M. D., Kusewitt, D., Mills, G. B., Kastan, M. B., & Walker, C. L. (2010). ATM signals to TSC2 in the cytoplasm to regulate mTORC1 in response to ROS. Proceedings of the National Academy of Sciences, 107(9), 4153–4158. 10.1073/pnas.0913860107

Bissler, J. J., & Kingswood, J. C. (2016). Optimal treatment of tuberous sclerosis complex associated renal angiomyolipomata: A systematic review. Therapeutic Advances in Urology, 8(4), 279–290. 10.1177/1756287216641353

Bottolo, L., Miller, S., & Johnson, S. R. (2020). Sphingolipid, fatty acid and phospholipid metabolites are associated with disease severity and mTOR inhibition in lymphangioleiomyomatosis. Thorax, 75(8), 679– 688. 10.1136/thoraxjnl-2019-214241

Buj Pradilla, M. J., Martí Ballesté, T., Torra, R., & Villacampa Aubá, F. (2017). Recommendations for imaging-based diagnosis and management of renal angiomyolipoma associated with tuberous sclerosis complex. Clinical Kidney Journal, 10(6), 728–737. 10.1093/ckj/sfx094

Carbonara, C., Longa, L., Grosso, E., Borrone, C., Garré, M. G., Brisigotti, M., & Migone, N. (1994). 9q34 loss of heterozygosity in a tuberous sclerosis astrocytoma suggests a growth suppressor-like activity also for the TSC1 gene. Human Molecular Genetics, 3(10), 1829–1832. 10.1093/hmg/3.10.1829

Chang, W. Y., Cane, J. L., Blakey, J. D., Kumaran, M., Pointon, K. S., & Johnson, S. R. (2012). Clinical utility of diagnostic guidelines and putative biomarkers in lymphangioleiomyomatosis. Respiratory Research, 13(1), 34. 10.1186/1465-9921-13-34

Cook, J. A., Oliver, K., Mueller, R. F., & Sampson, J. (1996). A cross sectional study of renal involvement in tuberous sclerosis. Journal of Medical Genetics, 33(6), 480–484. 10.1136/jmg.33.6.480

Dabora, S. L., Jozwiak, S., Franz, D. N., Roberts, P. S., Nieto, A., Chung, J., Choy, Y.-S., Reeve, M. P., Thiele, E., Egelhoff, J. C., Kasprzyk-Obara, J., Domanska-Pakiela, D., & Kwiatkowski, D. J. (2001). Mutational Analysis in a Cohort of 224 Tuberous Sclerosis Patients Indicates Increased Severity of TSC2, Compared with TSC1, Disease in Multiple Organs. American Journal of Human Genetics, 68(1), 64–80.

Eijkemans, M. J. C., Wal, W. van der, Reijnders, L. J., Roes, K. C. B., Doorn-Khosrovani, S. B. van W. van, Pelletier, C., Magestro, M., & Zonnenberg, B. (2015). Long-term Follow-up Assessing Renal Angiomyolipoma Treatment Patterns, Morbidity, and Mortality: An Observational Study in Tuberous Sclerosis Complex Patients in the Netherlands. American Journal of Kidney Diseases, 66(4), 638–645. 10.1053/j.ajkd.2015.05.016

Ewalt, D. H., Sheffield, E., Sparagana, S. P., Delgado, M. R., & Roach, E. S. (1998). Renal lesion growth in children with tuberous sclerosis complex. Journal of Urology, 160(1), 141–145. 10.1016/S0022-5347(01)63072-6

Green, A. J., Smith, M., & Yates, J. R. W. (1994). Loss of heterozygosity on chromosome 16p13.3 in hamartomas from tuberous sclerosis patients. Nature Genetics, 6(2), Article 2. 10.1038/ng0294-193

Hatano, T., & Egawa, S. (2020). Renal angiomyolipoma with tuberous sclerosis complex: How it differs from sporadic angiomyolipoma in both management and care. Asian Journal of Surgery, 43(10), 967– 972. 10.1016/j.asjsur.2019.12.008

Henske, E. P., Jóźwiak, S., Kingswood, J. C., Sampson, J. R., & Thiele, E. A. (2016). Tuberous sclerosis complex. Nature Reviews Disease Primers, 2(1), Article 1. 10.1038/nrdp.2016.35

Jo, D. S., Park, N. Y., & Cho, D.-H. (2020). Peroxisome quality control and dysregulated lipid metabolism in neurodegenerative diseases. Experimental & Molecular Medicine, 52(9), Article 9. 10.1038/s12276-020-00503-9

Kingswood, J. C., Belousova, E., Benedik, M. P., Carter, T., Cottin, V., Curatolo, P., Dahlin, M., D’ Amato, L., d’Augères, G. B., de Vries, P. J., Ferreira, J. C., Feucht, M., Fladrowski, C., Hertzberg, C., Jozwiak, S., Lawson, J. A., Macaya, A., Marques, R., Nabbout, R., … Jansen, A. C. (2019). Renal angiomyolipoma in patients with tuberous sclerosis complex: Findings from the TuberOus SClerosis registry to increase disease Awareness. Nephrology Dialysis Transplantation, 34(3), 502–508. 10.1093/ndt/gfy063

Lam, H. C., Siroky, B. J., & Henske, E. P. (2018). Renal disease in tuberous sclerosis complex: Pathogenesis and therapy. Nature Reviews Nephrology, 14(11), Article 11. 10.1038/s41581-018-0059-6

Lazarow, P. B., & De Duve, C. (1976). A fatty acyl-CoA oxidizing system in rat liver peroxisomes; enhancement by clofibrate, a hypolipidemic drug. Proceedings of the National Academy of Sciences, 73(6), 2043–2046. 10.1073/pnas.73.6.2043

Litwin, J. A., Völkl, A., Stachura, J., & Fahimi, H. D. (1988). Detection of peroxisomes in human liver and kidney fixed with formalin and embedded in paraffin: The use of catalase and lipid β-oxidation enzymes as immunocytochemical markers. The Histochemical Journal, 20(3), 165–173. 10.1007/BF01746680

Lu, Q., Zong, W., Zhang, M., Chen, Z., & Yang, Z. (2022). The Overlooked Transformation Mechanisms of VLCFAs: Peroxisomal β-Oxidation. Agriculture, 12(7), Article 7. 10.3390/agriculture12070947

Mandrekar, J. N. (2010). Receiver Operating Characteristic Curve in Diagnostic Test Assessment. Journal of Thoracic Oncology, 5(9), 1315–1316. 10.1097/JTO.0b013e3181ec173d

Martin, K. R., Zhou, W., Bowman, M. J., Shih, J., Au, K. S., Dittenhafer-Reed, K. E., Sisson, K. A., Koeman, J., Weisenberger, D. J., Cottingham, S. L., DeRoos, S. T., Devinsky, O., Winn, M. E., Cherniack, A. D., Shen, H., Northrup, H., Krueger, D. A., & MacKeigan, J. P. (2017). The genomic landscape of tuberous sclerosis complex. Nature Communications, 8(1), Article 1. 10.1038/ncomms15816

McCarthy, C., Gupta, N., Johnson, S. R., Yu, J. J., & McCormack, F. X. (2021). Lymphangioleiomyomatosis: Pathogenesis, clinical features, diagnosis, and management. The Lancet Respiratory Medicine, 9(11), 1313–1327. 10.1016/S2213-2600(21)00228-9

Nair, N., Chakraborty, R., Mahajan, Z., Sharma, A., Sethi, S. K., & Raina, R. (2020). Renal Manifestations of Tuberous Sclerosis Complex. Journal of Kidney Cancer and VHL, 7(3), 5–19. 10.15586/jkcvhl.2020.131

Nijmeh, J., El-Chemaly, S., & Henske, E. P. (2018). Emerging biomarkers of lymphangioleiomyomatosis. Expert Review of Respiratory Medicine, 12(2), 95–102. 10.1080/17476348.2018.1409622

Northrup, H., Aronow, M. E., Bebin, E. M., Bissler, J., Darling, T. N., de Vries, P. J., Frost, M. D., Fuchs, Z., Gosnell, E. S., Gupta, N., Jansen, A. C., Jóźwiak, S., Kingswood, J. C., Knilans, T. K., McCormack, F. X., Pounders, A., Roberds, S. L., Rodriguez-Buritica, D. F., Roth, J., … Young, L. (2021). Updated International Tuberous Sclerosis Complex Diagnostic Criteria and Surveillance and Management Recommendations. Pediatric Neurology, 123, 50–66. 10.1016/j.pediatrneurol.2021.07.011

O’Callaghan, F. J., Noakes, M. J., Martyn, C. N., & Osborne, J. P. (2004). An epidemiological study of renal pathology in tuberous sclerosis complex. BJU International, 94(6), 853–857. 10.1111/j.1464-410X.2004.05046.x

Polley, M.-Y. C., & Dignam, J. J. (2021). Statistical Considerations in the Evaluation of Continuous Biomarkers. Journal of Nuclear Medicine, 62(5), 605–611. 10.2967/jnumed.120.251520

Qiu, S., Cai, Y., Yao, H., Lin, C., Xie, Y., Tang, S., & Zhang, A. (2023). Small molecule metabolites: Discovery of biomarkers and therapeutic targets. Signal Transduction and Targeted Therapy, 8(1), Article 1. 10.1038/s41392-023-01399-3

Rakowski, S. K., Winterkorn, E. B., Paul, E., Steele, D. J. R., Halpern, E. F., & Thiele, E. A. (2006). Renal manifestations of tuberous sclerosis complex: Incidence, prognosis, and predictive factors. Kidney International, 70(10), 1777–1782. 10.1038/sj.ki.5001853

Reddy, J. K., & Hashimoto, T. (2001). PEROXISOMAL {beta}-OXIDATION AND PEROXISOME PROLIFERATOR-ACTIVATED RECEPTOR {{alpha}}: An Adaptive Metabolic System. Annual Review of Nutrition, 21(1), 193. 10.1146/annurev.nutr.21.1.193

Revilla-López, E., Ruiz de Miguel, V., López-Meseguer, M., Berastegui, C., Boada-Pérez, M., Mendoza-Valderrey, A., Arjona-Peris, M., Zapata-Ortega, M., Monforte, V., Bravo, C., Roman, A., Gómez-Ollés, S., & Sáez-Giménez, B. (2023). Lymphangioleiomyomatosis: Searching for potential biomarkers. Frontiers in Medicine, 10. https://www.frontiersin.org/articles/10.3389/fmed.2023.1079317

Roach, E. S., & Sparagana, S. P. (2004). Diagnosis of Tuberous Sclerosis Complex. Journal of Child Neurology, 19(9), 643–649. 10.1177/08830738040190090301

Sancak, O., Nellist, M., Goedbloed, M., Elfferich, P., Wouters, C., Maat-Kievit, A., Zonnenberg, B., Verhoef, S., Halley, D., & Van Den Ouweland, A. (2005). Mutational analysis of the TSC1 and TSC2 genes in a diagnostic setting: Genotype – phenotype correlations and comparison of diagnostic DNA techniques in Tuberous Sclerosis Complex. European Journal of Human Genetics, 13(6), 731–741. 10.1038/sj.ejhg.5201402

Seyama, K., Kumasaka, T., Souma, S., Sato, T., Kurihara, M., Mitani, K., Tominaga, S., & Fukuchi, Y. (2006). Vascular Endothelial Growth Factor-D Is Increased in Serum of Patients with Lymphangioleiomyomatosis. Lymphatic Research and Biology, 4(3), 143–152. 10.1089/lrb.2006.4.143

Shepherd, C. W., Gomez, M. R., Lie, J. T., & Crowson, C. S. (1991). Causes of Death in Patients With Tuberous Sclerosis. Mayo Clinic Proceedings, 66(8), 792–796. 10.1016/S0025-6196(12)61196-3

Titorenko, V. I., & Rachubinski, R. A. (2001). The life cycle of the peroxisome. Nature Reviews Molecular Cell Biology, 2(5), Article 5. 10.1038/35073063

Trnka, P., & Kennedy, S. E. (2021). Renal tumors in tuberous sclerosis complex. Pediatric Nephrology, 36(6), 1427–1438. 10.1007/s00467-020-04775-1

Walker, C. L., Pomatto, L. C. D., Tripathi, D. N., & Davies, K. J. A. (2018). Redox Regulation of Homeostasis and Proteostasis in Peroxisomes. Physiological Reviews, 98(1), 89–115. 10.1152/physrev.00033.2016

Wang, Z., Liu, X., Wang, W., Xu, J., Sun, H., Wei, J., Yu, Y., Zhao, Y., Wang, X., Liao, Z., Sun, W., Jia, L., & Zhang, Y. (2023). UPLC-MS based integrated plasma proteomic and metabolomic profiling of TSC-RAML and its relationship with everolimus treatment. Frontiers in Molecular Biosciences, 10, 1000248. 10.3389/fmolb.2023.1000248

Young, L. R., Inoue, Y., & McCormack, F. X. (2008). Diagnostic Potential of Serum VEGF-D for Lymphangioleiomyomatosis. New England Journal of Medicine, 358(2), 199–200. 10.1056/NEJMc0707517

Zhang, J., Kim, J., Alexander, A., Cai, S., Tripathi, D. N., Dere, R., Tee, A. R., Tait-Mulder, J., Di Nardo, A., Han, J. M., Kwiatkowski, E., Dunlop, E. A., Dodd, K. M., Folkerth, R. D., Faust, P. L., Kastan, M. B., Sahin, M., & Walker, C. L. (2013). A tuberous sclerosis complex signalling node at the peroxisome regulates mTORC1 and autophagy in response to ROS. Nature Cell Biology, 15(10), Article 10. 10.1038/ncb2822

Zhang, J., Tripathi, D. N., Jing, J., Alexander, A., Kim, J., Powell, R. T., Dere, R., Tait-Mulder, J., Lee, J.-H., Paull, T. T., Pandita, R. K., Charaka, V. K., Pandita, T. K., Kastan, M. B., & Walker, C. L. (2015). ATM functions at the peroxisome to induce pexophagy in response to ROS. Nature Cell Biology, 17(10), Article 10. 10.1038/ncb3230

TSC Natural History Database. Available from: https://www.tscalliance.org/get-involved/participate-in-research/tsc-biosample-repository-and-natural-history-database/

